# High-throughput extraction of SARS-CoV-2 RNA from nasopharyngeal swabs using solid-phase reverse immobilization beads

**DOI:** 10.1101/2020.04.08.20055731

**Authors:** Mahesh K. R. Kalikiri, Mohammad Rubayet Hasan, Faheem Mirza, Thabisile Xaba, Patrick Tang, Stephan Lorenz

## Abstract

The ongoing pandemic of the novel coronavirus, SARS-CoV-2, has led to a global surge in laboratory testing for the virus. The gold standard approach to detecting an active viral infection is the use of RT-qPCR. This approach requires the isolation of viral RNA from respiratory specimens, such as nasopharyngeal swabs.

We developed a method using a widely available lysis buffer coupled with solid-phase reverse immobilization (SPRI) beads to extract viral RNA from swabs collected in viral transport medium (VTM) which can be performed manually or on a Hamilton STAR liquid-handling robot. Using a WHO recommended, laboratory-developed RT-qPCR for SARS-CoV-2, we validated this method in a CAP-accredited laboratory, against the IVD-labelled bioMérieux NucliSENS easyMAG automated extraction platform.

Our method demonstrates a comparable sensitivity and specificity, making it suitable for large-scale testing and monitoring of suspected COVID-19 cases and health care workers. This is especially important as the world faces critical shortages of viral RNA extraction reagents for the existing commercial extraction systems.

## INTRODUCTION

In December 2019, the first reports of a new pathogenic virus causing severe respiratory illness emerged (1). Early investigations determined that this new disease was caused by a novel coronavirus, SARS-CoV-2. Due to the rapid transmission and spread of the disease, the WHO declared this outbreak a global pandemic on 31^st^ January 2020 (2). Because of the wide spectrum of presentations, from asymptomatic and subclinical infections to pneumonia and acute respiratory distress syndrome, many of which resemble other common respiratory infections, laboratory testing is essential for establishing the diagnosis of COVID-19. Once infected individuals are identified through laboratory testing, they can be isolated and managed appropriately, along with identification and quarantine of their contacts. The increased demand for laboratory testing has led to global shortages of all the required supplies, from personal protective equipment such as N95 masks, to swabs, and even the reagents required for viral RNA extraction.

Today, most laboratories performing extraction of RNA from samples rely heavily on commercially available kits due to their ease of use and standardization across laboratories. When these kits become scarce, many laboratories will have to turn to alternative methods, but often lack the expertise or the resource to test and validate those alternative methods. The underlying principle of modern, paramagnetic bead-based isolation of nucleic acids is their precipitation from solution by a molecular crowding reagent such as polethylenglycol (PEG) under high salt conditions. It was first described for DNA 45 years ago (1) and later adapted to the isolation of viral RNA from tissues (2). Modern iterations of this procedure such as Ampure XP add carboxyl-coated paramagnetic beads to the PEG solution, thus greatly accelerating the precipitation as well as alleviating the need for time-consuming centrifugations, enabling automation of this process on liquid handling robots (3). This method is highly efficient in recovering even minute amounts of nucleic acids from larger volumes, such as shown in the precipitation of single-cell genomes (4,5).

Here, we exploited the use of paramagnetic beads and readily available lysis buffers in combination with a RT-qPCR assay based on the SARS-CoV-2 E gene assay, first described by Corman et al. (6). The method presented here provides a means to extract RNA from swabs of suspected COVID-19 cases that can be readily implemented in many laboratories, without relying on scarce extraction kits.

## RESULTS

### RLT buffer used in 1:1 ratio with VTM is efficient at lysing respiratory viruses

Retrospective, residual nasopharyngeal specimens that previously tested positive or negative for respiratory syncytial virus (RSV) using the Cepheid GeneXpert Flu/RSV kit or the Fast Track Diagnostics Respiratory Pathogens 21 PCR (FTD-RESP21) were extracted with our new approach on a Hamilton STAR liquid handling robot with 8 CO-RE channels. In brief, we added 150 µl of buffer RLT to 150 µl of viral transport medium for inactivation of the virus. We precipitated the contained RNA using 300 µl of Ampure XP beads, performed an extensive wash and eluted the RNA in 20 µl. For comparison, the same samples were extracted using the IVD-labelled bioMérieux NucliSENS easyMAG automated extraction platform, using 200 µl of VTM and eluted in 40 µl. We used 5 µl of the eluates of both extraction methods in our in-house LDT-RSV RT-qPCR assay.

Across 47 total samples tested, the extraction method had 91% sensitivity, 100% specificity and 96% accuracy (Table 1) with 2 false negatives. All results produced by the new method were within <2 Cq of the validated in-house protocol (7). Two samples that gave false negative results had Cq values ≥35, suggesting that the slightly lower sample volume that is fed into the extraction leads to slightly reduced sensitivity.

**Table 1:** Detection of RSV as surrogate marker for SARS-CoV-2. RSV-positive samples were extracted with the new method and compared against the reference method.

| Statistics | Value | 95% CI |
| --- | --- | --- |
| Total number of results | 47 | - |
| True positive | 21 | - |
| True negative | 24 | - |
| False positive | 0 | - |
| False negative | 2 | - |
| Sensitivity | 91% | 72-99% |
| Specificity | 100% | 86-100% |
| Accuracy | 96% | 85-99% |

### Addition of carrier RNA increases recovery of viral RNA

Various manufacturers supplement their extraction kits with non-specific carrier RNA at high amounts (up to 3 µg, Qiagen) to create a crowding or “pull-down” effect in samples with low endogenous amounts of RNA. It is also speculated that carrier RNA act as a protectant against RNAses contained in biological samples.

We extracted RNA from freshly harvested MS2 phage spiked into 150 µl of VTM in the presence of 0 and 1 µg carrier RNA (Qiagen) and observed an increase of the amount of MS2 RNA detected (Figure 1), as well as a reduced variation between samples, when extracted on our liquid-handling system.

**Figure 1:**
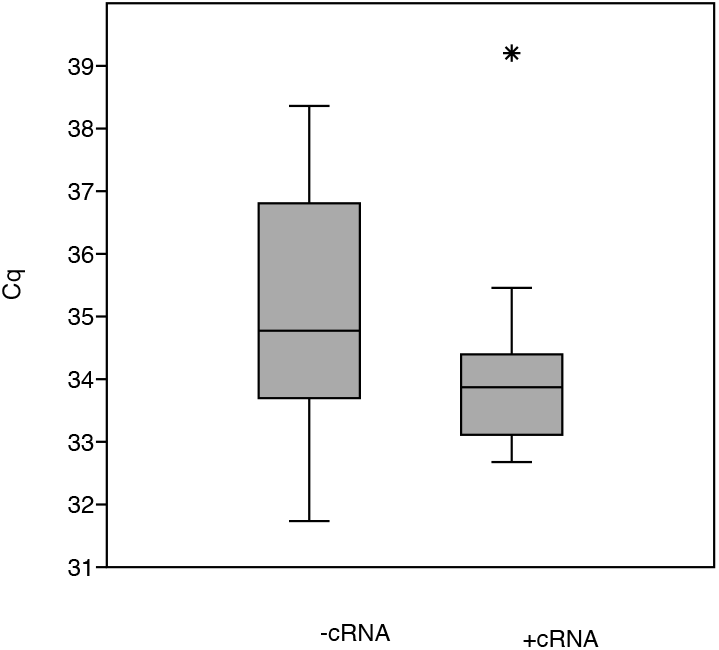
Phage MS2 RNA recovery improves by adding carrier RNA. Without carrier RNA (-cRNA), 10 out of 24 samples failed entirely and the detected samples show a Cq of 35.2 ± 0.51SE. Addition of 1 µg carrier RNA to the extraction (+cRNA) leads to a positive detection in 22 out of 24 samples with an average Cq of 34.0 ± 0.28 SE.

When adding between 0 and 1 µg carrier RNA in 0.25 µg increments directly into the one-step RT-qPCR reaction assay detecting a SARS-CoV-2 positive control, we observed a decrease in Cq directly correlated to the amount of carrier RNA (Pearson r^=^0.985, Figure 2) present in the reaction. For all subsequent experiments we added 0.5 µg of carrier RNA per extraction sample, mixed into the RLT buffer.

**Figure 2:**
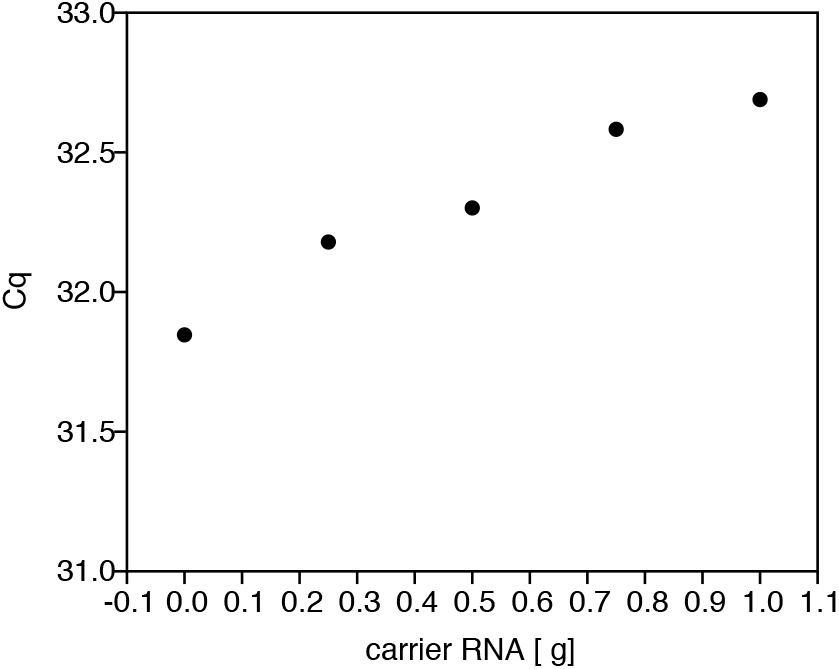
Detection of SARS-CoV-2 positive control is suppressed carrier RNA. RT-qPCR was performed on duplicate positive control samples with increasing amounts of carrier RNA spiked into the reaction (x-axis). Addition of 1 µg of carrier RNA leads to a loss of 0.84 Cq (y-axis).

The sensitivity of the assay was further increased by extending the drying step of the extraction protocol. After increasing the drying step to at least 7 min, no inhibition of MS2 detection was observed.

### RLT lysis followed by automated bead capture and washing yield comparable results with SARS-CoV-2 to standard extraction method

After establishing the protocol and implementing a version extracting 96 samples at a time on a Hamilton STAR, we tested 94 clinical samples side-by-side using the Biomérieux easyMAG system and the proposed method (Table 2). Apart from one sample that tested weakly positive at Cq 38 for the validated method, all positive samples were correctly identified, with a Cq difference of <1 Cq. More importantly, only 3 out of 94 samples exhibited PCR inhibition as determined by the successful detection of the internal control. Overall, the PCR inhibition rate was 3.2% for the proposed method, compared with 11.2% for the established commercial method.

**Table 2:** Detection of SARS-CoV-2 from nasopharyngeal swabs extracted with the novel method compared to the reference method.

| Statistics | Value | 95% CI |
| --- | --- | --- |
| Total number of results | 94 | - |
| True positive | 14 | - |
| True negative | 79 | - |
| False positive | 0 | - |
| False negative | 1 | - |
| Sensitivity | 93% | 68-100% |
| Specificity | 100% | 95-100 |
| Accuracy | 99% | 94-100 |

We also tested another subset of 204 pooled clinical samples (5 samples per pool), of which 69 (33.8%) tested positive, 130 (63.7%) negative, and 5 (2.5%) showed signs of PCR inhibition.

## DISCUSSION

Many of the commercially available kits for the extraction of RNA from samples are based on long-standing and well-known principles. Our results show that the commonly available buffer RLT can lyse RSV, SARS-CoV-2 and bacteriophage MS2 reliably when used in a 1:1 ratio.

The precipitation effect of PEG solution and size of precipitated fragments is driven by the ratio of PEG to the nucleic acid solution. Therefore, we established this method with a relatively high 1:1 ratio of Ampure XP beads to lysed sample, which is known to precipitate fragments larger than 200 bp. While this approach is more reagent consuming, it guarantees the capture of even small fragments of viral RNA from potentially degraded samples and is likely a contributing factor to the observed sensitivity. Future iterations of this protocol could focus on cost-reduction by minimizing the amount of costly Ampure XP beads by substituting parts of the volume with ethanol or isopropanol, which do not show a ratio-dependent size-selection of nucleic acids.

Our results show that the addition of carrier RNA is a double-edged sword. While the addition increases the extraction efficiency and consistency, we hypothesized that an excess of carrier RNA may inhibit the reverse transcription. Indeed, we observed a small negative effect of carrier RNA on the detection of a transcript when directly added to the RT-qPCR reaction. Until more experiments are done to fully understand the intricacies of this effect, we opted for a compromise between increased capture efficiency and consistency and a weak suppressive effect on the reverse transcription efficiency. When applying these conditions to 204 clinical samples, we only observed a 2.5% inhibition of the RT-qPCR, indicating that our method is suitable for large-scale use in testing laboratories.

## CONCLUSIONS

In this manuscript, we demonstrate the feasibility of using reagents commonly available in molecular biology and next-generation sequencing laboratories for the extraction of SARS-CoV-2 viral RNA for the diagnosis of COVID-19. This method has a comparable sensitivity and specificity with commercially available reference assays, especially when considering that the reference method uses 220 µl of sample, whereas this method only uses 150 µl. This method can be performed manually, with a throughput of maximum 48 samples in 90 minutes. In order to increase throughput and reproducibility and reduce human error, it can be implemented on many laboratory liquid-handling robots. In our hands, we were able to process 96 samples in 60 minutes. We made 2 methods for a Hamilton STAR liquid-handling system available on GitHub, processing either 24 or 96 samples per run. Future versions will make use of on-deck parallelization and offer a throughput of at least 192 samples in 60 min.

## METHODS

### RNA extraction using SPRI beads

150 µl of VTM specimens were mixed with an equal volume of buffer RLT (Qiagen) containing 6.7 µg/µl carrier RNA (Qiagen) and incubated at RT for 5 min. 300 µl of AmpureXP paramagnetic beads (Beckman Coulter) were added to the sample and incubated for 6 min while mixing, either with a pipette or on a shaker at 1,600 rpm. The beads were captured by incubating the plate for 8-10 min. The supernatant was discarded, and the bead pellet washed twice with 300 µl of freshly prepared 80% ethanol. During the first wash, the pellet was resuspended and incubated on the magnet for 6 minutes. No mixing was performed during the second ethanol wash. The pellet was air-dried and the absence of ethanol visually confirmed. The RNA was eluted by adding 23 µl of RSB (Illumina) and transferring 20 µl of eluate to a fresh plate.

### RT-qPCR detection of RSV

RT-qPCR assay was performed using a laboratory-developed test as described previously (7).

### RT-qPCR detection of SARS-CoV-2

We used the QuantiFast Pathogen +IC (Qiagen) kit for the detection of the SARS-CoV-2 E gene and the internal control on a 7500 Fast RT-PCR instrument (Applied Biosystems). The master mix was created using 2.5 µl 5x QuantiFast Pathogen master mix, 0.1 µl 100x QuantiFast RT mix,

0.25 µl 50x ROX dye solution and 0.9 µl RNAse-free water and 3.5 µl primer-probe mix (to final concentration of 0.2 µM and 0.4 µM, respecitvely). The following primers and concentrations were used:

E_Sarbeco-F1: 5’–ACAGGTACGTTAATAGTTAATAGCGT-3’ (0.4 µM)

E-Sarbeco-R2: 5’-ATATTGCAGCAGTACGCACACA-3’ (0.4 µM)

E-Sarbeco-P1: 5’-FAM/ZEN-ACACTAGCCATCCTTACTGCGCTTCG-IaBkFQ-3’

(0.2 µM)

7.5 µl of master mix were mixed with 5 µl of extracted RNA and incubated at 50°C for 20 min, followed by 5 min at 95°C and 45 cycles of 15 s at 95°C and 30 s at 60°C.

## Data Availability

Raw data will be made available on request. Supplementary package files to run this method on a Hamilton STAR liquid handler can be found on https://github.com/Sidra-IGS/covid-19

## ACKNOWLEDGEMENTS

COVID-19 samples were collected and referred for testing by Hamad Medical Corporation. We would like to thank Dr. Khalid Fakhro, Sidra Medicine, for his valuable input and discussion.

## CONTRIBUTIONS

MK performed the method development experiments and implemented the method on the automation platform. MR performed additional tests, RT-qPCR data analysis and test design. FM & TX prepared and tested the clinical samples during the validation. PT contributed to the design of the method and writing of the manuscript. SL designed the proof of concept, designed tests and automation concepts and wrote the manuscript.

**The authors declare no conflict of interest.**

## SUPPLEMENTARY MATERIAL

The protocol for a Hamilton STAR liquid-handling robot processing either 24 or 96 samples at a time, alongside an SOP, can be found at https://github.com/Sidra-IGS/covid-19.

The detailed protocol can be found in supplement 1.

## REFERENCES

1. Lis, J. T. & Schleif, R. Size fractionation of double-stranded DNA by precipitation with polyethylene glycol. Nucleic Acids Res. 2, 383–389 (1975).

2. Yang, F. & Xu, X. A new method of RNA preparation for detection of hepatitis A virus in environmental samples by the polymerase chain reaction. J. Virol. Methods 43, 77–84 (1993).

3. DeAngelis, M. M., Wang, D. G. & Hawkins, T. L. Solid-phase reversible immobilization for the isolation of PCR products. Nucleic Acids Res. 23, 4742–4743 (1995).

4. Macaulay, I. C. et al. Separation and parallel sequencing of the genomes and transcriptomes of single cells using G&T-seq. Nat. Protoc. 11, 2081–2103 (2016).

5. Bronner, I. F. & Lorenz, S. Combined Genome and Transcriptome (G&T) Sequencing of Single Cells. Methods Mol. Biol. Clifton NJ 1979, 319–362 (2019).

6. Corman, V. M. et al. Detection of 2019 novel coronavirus (2019-nCoV) by real-time RT-PCR. Eurosurveillance 25, (2020).

7. Hasan, M. R. et al. A novel real-time PCR assay panel for detection of common respiratory pathogens in a convenient, strip-tube array format. J. Virol. Methods 265, 42–48 (2019).

